# Retrospective Study of Bacteremia Caused by *Alistipes* Species in Japan, 2016–2023

**DOI:** 10.1101/2024.08.24.24312474

**Authors:** Naoki Watanabe, Tomohisa Watari, Naoto Hosokawa, Yoshihito Otsuka

**Author notes:** Corresponding Author: Naoki Watanabe Kameda Medical Center, 296-0044, Higashi-cho 929, Kamogawa, Chiba, Japan.

## Abstract

The clinical characteristics of *Alistipes* bacteremia remain insufficiently described. Therefore, here, we retrospectively analyzed 13 cases of *Alistipes* bacteremia at a tertiary care center to evaluate their clinical and microbiological characteristics. Ten patients were aged ≥ 65 years, and seven were female. Nine patients had comorbidities, including seven with solid tumors or hematological malignancies. Eleven patients presented with gastrointestinal symptoms. The isolates included four *Alistipes finegoldii*, four *Alistipes onderdonkii*, three *Alistipes putredinis*, two *Alistipes indistinctus*, and one *Alistipes ihumii* isolates. Ten available strains exhibited low minimum inhibitory concentrations against β-lactam/β-lactamase inhibitors and metronidazole, whereas they exhibited high minimum inhibitory concentrations for penicillin, ceftriaxone, and minocycline. Several strains harbored antimicrobial resistance genes, including *adeF*, *tet*(Q), *cfxA3*, *cfxA4*, and *erm*(G). Twelve patients received β-lactam/β-lactamase inhibitors, and two elderly patients with solid tumors developed septic shock and died. *Alistipes* species can translocate from the gastrointestinal tract into the bloodstream, leading to severe infections.

## Introduction

*Alistipes* is a rarely detected anaerobic bacterium in clinical samples. Initially described as a bile-resistant, pigment-producing, Gram-negative bacillus (*1*, *2*), *Alistipes* was classified as a new genus in 2003 (*3*). The genus *Alistipes* includes 11 species, including *Alistipes finegoldii* (*3*), *Alistipes putredinis* (*3*), and *Alistipes onderdonkii* (*4*). These species have been isolated from the appendix tissue, feces, intra-abdominal fluid, and blood (*3*,*5–7*). Although the role of *Alistipes* in humans remains unclear, this bacterium may play a preventive or pathogenic role in various diseases. *Alistipes* may help to prevent colitis or contribute to the pathogenesis of depression (*8*, *9*).

Anaerobic bacteria, which are a part of the normal human flora, are also significant pathogens in bacteremia and other infectious diseases. Anaerobic bacteremia is often associated with abdominal, pelvic, skin, and soft tissue infections (*10*). *Alistipes* is associated with infections, such as appendicitis (*1*, 2), and intra-abdominal infections (*11*). Three cases of *Alistipes* bacteremia, with patients experiencing colon cancer (*12*) or peritonitis (*13*), have been reported.

Owing to the limited number of reports, the clinical characteristics of *Alistipes* bacteremia remain insufficiently described.

Inadequate treatment of anaerobic bacterial infections can lead to clinical failure. In cases of anaerobic bacteremia, the mortality rate is higher in patients receiving ineffective antimicrobial therapy than in those receiving appropriate antimicrobial therapy (*14*). Therefore, understanding the antimicrobial susceptibility of *Alistipes* species is crucial for implementing effective treatments. Of the three *Alistipes* strains isolated from the blood, two of them have shown low susceptibility to penicillin (PEN) and one to cefotetan (*12*, *13*). Currently, the antimicrobial susceptibility of *Alistipes* isolates from the blood is not well understood.

Therefore, this study aimed to investigate and describe the clinical characteristics of *Alistipes* bacteremia in a tertiary care center in Japan. In addition, we evaluated the accuracy of identification testing for *Alistipes* species and assessed the antimicrobial resistance of the isolates.

## Materials and Methods

### Study Design and Data Collection

This was a retrospective cohort study of *Alistipes* bacteremia at Kameda Medical Center in Japan conducted between April 2016 and December 2023. We reviewed 31,875 patients who underwent blood cultures and identified *Alistipes* cases. For patients with multiple episodes, only the first episode was included in this study. Patient data and microbiological testing results were collected from medical records and a laboratory system. Patient data included age, sex (male or female), infection type (community-acquired, healthcare-associated), comorbidities, symptoms, infection sites, procedures, antimicrobial therapy, hospitalization days from blood collection, and outcomes. Healthcare-associated infections were defined as cases with one of the following conditions: onset after 48 h of hospitalization or patients receiving medical care, such as a nursing home, dialysis, or outpatient chemotherapy. The Ethics Committee of Kameda Medical Center approved this study (agreement no. 23-121). Written informed consent was not required because the data used were anonymized.

### Blood Culture and Identification Testing

Blood cultures were performed using BD Bactec™ FX Blood Culture System (Becton, Dickinson and Company, https://www.bd.com). Blood samples were collected using Plus Aerobic and Lytic Anaerobic Media for adult patients and Peds Plus Medium for pediatric patients (Becton, Dickinson and Company). The blood cultures were incubated at 35 °C for 7 days. The number of positive blood culture sets, time to positivity, and detected *Alistipes* were recorded. *Alistipes* strains were stored at –80 °C for identification, whole genome sequencing, and antimicrobial susceptibility testing. The preserved strains were cultured anaerobically using Brucella HK agar medium (Kyokuto Pharmaceutical Industrial, https://www.kyokutoseiyaku.co.jp) at 35 °C for 2 days and were used for each test.

The strains were identified using matrix-assisted laser desorption/ionization time-of-flight mass spectrometry (MALDI-TOF MS). MALDI-TOF MS was performed using the MALDI Biotyper system, which included the microflex LT/SH and flexControl version 3.4 (Bruker Daltonics GmbH, https://www.bruker.com). *Alistipes* colonies that developed on the Brucella HK agar medium were selected and applied to the test plate. Mass spectra were analyzed using the MALDI Biotyper Library, which recorded the highest score for the predicted species.

Identification results were accepted at the species level for scores of ≥ 2.0. In cases where an identification result could not be obtained using the standard cell smear method, the strains were retested using the formic acid on-plate or formic acid extraction method.

### Polymerase Chain Reaction (PCR) and Whole Genome Analysis

The identification results were validated by 16S rRNA gene and whole genome analyses of the preserved strains. Non-preserved strains were excluded from this analysis. Genomic DNA was extracted from the strains using magLEAD and Consumable Kits (Precision System Science; https://www.pss.co.jp). GeneAtlas Type G (Astec, https://www.astec-corp.co.jp) was used as the thermal cycler and Premix Taq Hot Start Version (Takara Bio, https://www.takara-bio.com) for the PCR reagent. Using previously reported primers for the 16S rRNA gene (*15*, *16*), PCR was performed according to the literature (*17*). PCR conditions and primers used are listed in Table S1. Sequencing was performed using FASMAC (https://fasmac.co.jp), and the resulting sequences were analyzed using EzBioCloud (*18*) for species identification. Additionally, 16S rRNA gene reanalysis was performed on three *A. finegoldii* strains (3302398, 4401054, and Granada) from previous bacteremia cases (*12*, *13*). The sequence data for 3302398 (accession no. AY547271), 4401054 (AY643082), and Granada (OM900033) were obtained from GenBank and analyzed using the same methods as the strains collected in this study.

Fragment libraries for the whole genome analysis were prepared using Illumina DNA Prep Tagmentation (M) Beads (Illumina, https://www.illumina.com). Sequencing was performed using the MiSeq Reagent Micro Kit (Illumina) to generate paired-end reads. De novo assembly was conducted using the CLC Genomics Workbench version 22.0.2 (Qiagen, https://www.qiagen.com). CheckM (*19*) was used as a quality check for the genome, and samples with a contamination rate of ≥ 5% were excluded from subsequent analysis. The average nucleotide identity (ANI) was performed using DFAST (*20*), identifying bacterial species with a threshold of ≥ 95%. For subspecies analysis, DNA-DNA hybridization (DDH) analysis was performed using the Genome–Genome Distance Calculator 3.0 (https://ggdc.dsmz.de) (*21*). The threshold for defining subspecies was set at 79% DDH of the previously reported criteria for subspecies classification (*22*). The presence of antimicrobial resistance genes was analyzed using the Comprehensive Antibiotic Resistance Database (*23*).

### Antimicrobial Susceptibility Testing

The antimicrobial susceptibility of the preserved strains was evaluated using the broth microdilution method with Brucella Broth and Dry Plates (Eiken Chemical, https://www.eiken.co.jp). The inoculum was prepared according to the manufacturer’s instructions and transferred to a microplate at a final volume of approximately 1 × 10^5^ colony-forming unit per well. After inoculation, the microplate was incubated anaerobically at 35–37 °C for 46–48 h. Minimum inhibitory concentrations (MICs) were determined after confirming bacterial growth in the control wells. The Dry Plate included the following antibiotics: PEN, ampicillin-sulbactam (SAM), amoxicillin-clavulanic acid (AMC), piperacillin-tazobactam (TZP), ceftriaxone (CRO), cefoxitin (FOX), imipenem (IPM), clindamycin (CLI), minocycline (MINO), moxifloxacin (MXF), and metronidazole (MTZ). After determining the MICs, the concentrations required to inhibit 50% and 90% of the strains (MIC_50_ and MIC_90_) were calculated.

### Statistical Analyses

The characteristics of *Alistipes* cases were summarized using EZR version 1.54 (*24*). Continuous variables are reported as medians and interquartile ranges (IQRs), and categorical variables are reported as actual numbers. The proportion of patients with *Alistipes* bacteremia among all patients who underwent blood culture was calculated using 95% confidence intervals. Time to positivity was reported as the median and IQR, excluding the results from blood culture sets with multiple pathogens.

## Results

### Patient Characteristics and Clinical Courses

Thirteen *Alistipes* bacteremia were identified over a 7-year period. This number represented 0.04% of the patients with blood cultures, and all cases were single episodes. The characteristics of the patients with *Alistipes* bacteremia are shown in Table 1. The median patient age was 72 (IQR, 65–85) years, and 10 patients were aged > 65 years. Seven patients were female, and nine patients had comorbid conditions, with a Charlson score ≥ 1. Six patients had solid tumors, and one had a hematological malignancy. The most common clinical symptom was abdominal pain (nine patients), followed by fever (six patients). Of the 13 patients, 11 presented with gastrointestinal symptoms, including abdominal pain, vomiting, diarrhea, and/or abdominal distention. The median time to positivity was 81 (IQR, 71–106) h, and *Alistipes* strains from three cases were detected in blood cultures that tested positive ≥ 120 h after the start of incubation. Four patients had two sets of positive blood cultures.

**Table 1.** Patient background of *Alistipes* bacteremia in a tertiary care center in Japan, 2016–2023

| Characteristics | All (n = 13) | Alive (n = 11) | Death (n = 2) |
| --- | --- | --- | --- |
| Age, median years (IQR) | 72 (65–85) | 70 (59–85) | 80 (78–83) |
| ≥ 65 years, no. (%) | 10 (76.9) | 8 (72.7) | 2 (100.0) |
| Sex, F, no. (%) | 7 (53.8) | 6 (54.5) | 1 (50.0) |
| Site of acquisition, no. (%) |  |  |  |
| Community | 7 (53.8) | 7 (63.6) | 0 (0.0) |
| Healthcare-associated | 6 (46.2) | 4 (36.4) | 2 (100.0) |
| Charlson score, no. (%) |  |  |  |
| 0 | 4 (30.8) | 4 (36.4) | 0 (0.0) |
| 1–2 | 4 (30.8) | 3 (27.3) | 1 (50.0) |
| 3–4 | 2 (15.4) | 2 (18.2) | 0 (0.0) |
| ≥ 5 | 3 (23.1) | 2 (18.2) | 1 (50.0) |
| Comorbid conditions, no. (%) |  |  |  |
| Solid tumor | 6 (46.2) | 4 (36.4) | 2 (100.0) |
| Hematological malignancies | 1 (7.7) | 1 (9.1) | 0 (0.0) |
| Leukopenia | 2 (15.4) | 2 (18.2) | 0 (0.0) |
| Chemotherapy | 3 (23.1) | 3 (27.3) | 0 (0.0) |
| Immunosuppressive therapy | 0 (0.0) | 0 (0.0) | 0 (0.0) |
| Intravascular device, no. (%) | 3 (23.1) | 2 (18.2) | 1 (50.0) |
| Symptoms, no. (%) |  |  |  |
| Abdominal pain | 9 (69.2) | 8 (72.7) | 1 (50.0) |
| Diarrhea | 1 (7.7) | 1 (9.1) | 0 (0.0) |
| Sepsis or septic shock | 3 (23.1) | 1 (9.1) | 2 (100.0) |
| Fever | 6 (46.2) | 6 (54.5) | 0 (0.0) |
| Vomiting | 3 (23.1) | 3 (27.3) | 0 (0.0) |
| Time to positivity, h (IQR) | 81 (71–106) | 76 (68–103) | 94 (88–99) |

The clinical presentations, treatments, and outcomes of *Alistipes* bacteremia are summarized in Table 2. Eleven of thirteen patients were diagnosed with gastrointestinal tract disease, with infection sites in these areas. The remaining two patients had *Alistipes* bacteremia with unknown infection sites (cases 7 and 12). The patient in case 7 developed a fever during treatment for a catheter-related bloodstream infection caused by *Staphylococcus aureus*, and *Alistipes indistinctus* was detected in a blood culture collected at the time of fever. The patient in case 12 had a history of a cerebral infarction and cholecystectomy. The patient presented with chills, fever, and vomiting, and blood cultures were positive for *A. putredinis*. All patients were clinically diagnosed with true bacteremia and received antimicrobial therapy. The duration of the antimicrobial therapy ranged from 13 to 36 days. The most commonly prescribed antimicrobials were β-lactam/β-lactamase inhibitors (BLBLIs), with nine patients receiving SAM and six patients receiving TZP. Six patients were positive for multiple pathogens, including *Escherichia coli*, *Klebsiella pneumoniae*, *Bacteroides* species, and others.

**Table 2.**
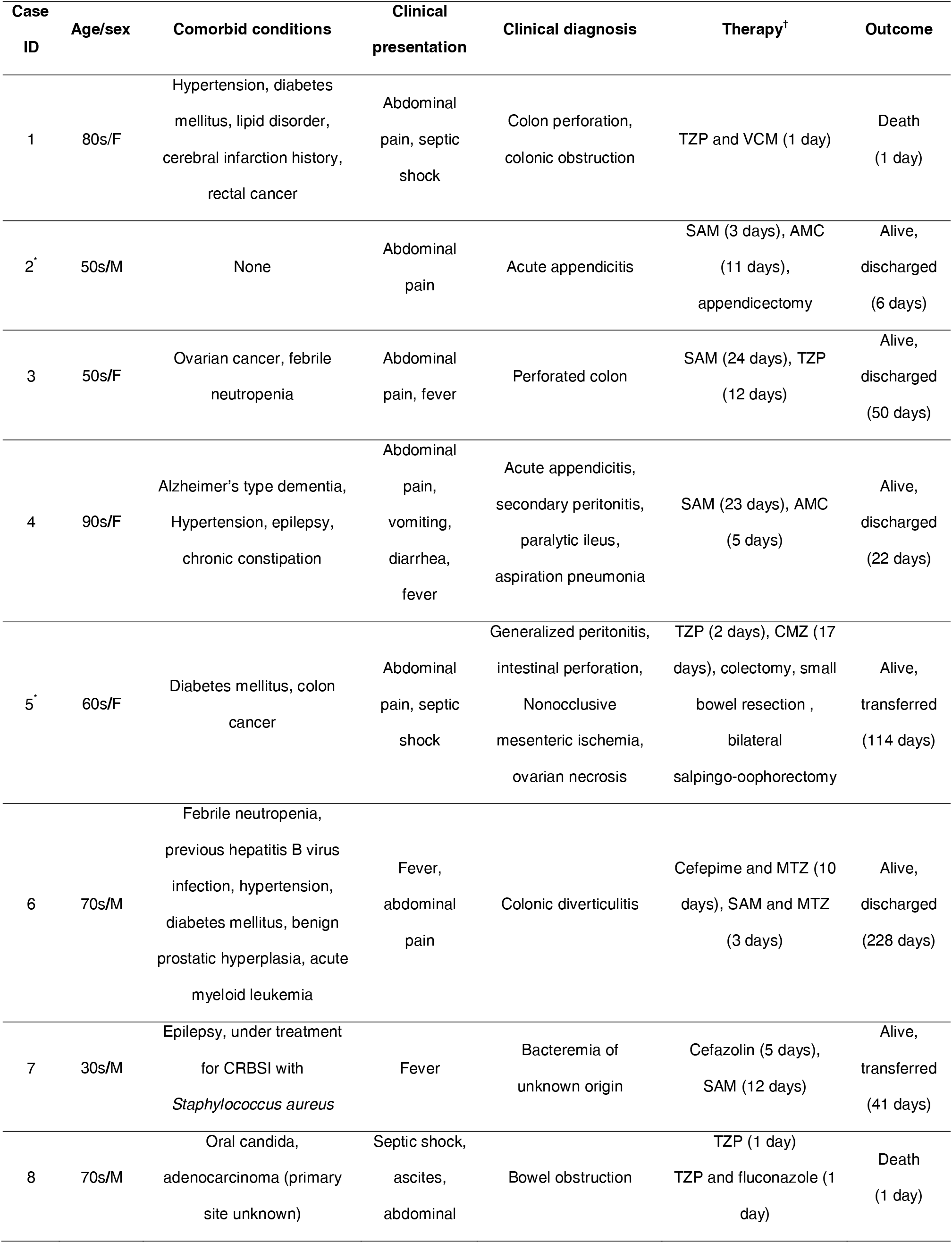
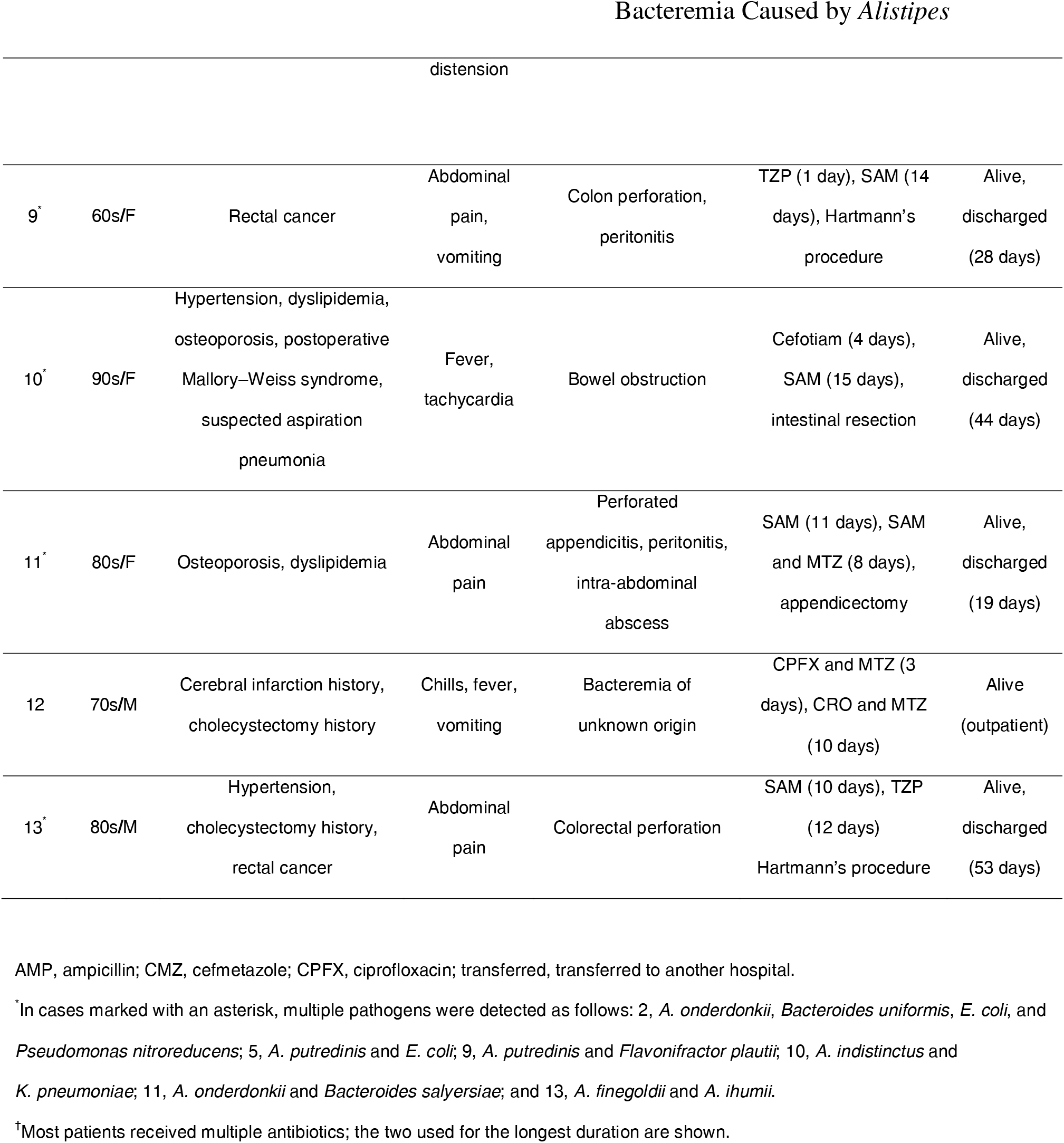
Patient characteristics and clinical course of *Alistipes* bacteremia in a tertiary care center in Japan, 2016–2023

| Case ID | Age/sex | Comorbid conditions | Clinical presentation | Clinical diagnosis | Therapy<sup>†</sup> | Outcome |
| --- | --- | --- | --- | --- | --- | --- |
| 1 | 80s/F | Hypertension, diabetes mellitus, lipid disorder, cerebral infarction history, rectal cancer | Abdominal pain, septic shock | Colon perforation, colonic obstruction | TZP and VCM (1 day) | Death (1 day) |
| 2<sup>*</sup> | 50s/M | None | Abdominal pain | Acute appendicitis | SAM (3 days), AMC (11 days), appendicectomy | Alive, discharged (6 days) |
| 3 | 50s/F | Ovarian cancer, febrile neutropenia | Abdominal pain, fever | Perforated colon | SAM (24 days), TZP (12 days) | Alive, discharged (50 days) |
| 4 | 90s/F | Alzheimer's type dementia, Hypertension, epilepsy, chronic constipation | Abdominal pain, vomiting, diarrhea, fever | Acute appendicitis, secondary peritonitis, paralytic ileus, aspiration pneumonia | SAM (23 days), AMC (5 days) | Alive, discharged (22 days) |
| 5<sup>*</sup> | 60s/F | Diabetes mellitus, colon cancer | Abdominal pain, septic shock | Generalized peritonitis, intestinal perforation, Nonocclusive mesenteric ischemia, ovarian necrosis | TZP (2 days), CMZ (17 days), colectomy, small bowel resection, bilateral salpingo-oophorectomy | Alive, transferred (114 days) |
| 6 | 70s/M | Febrile neutropenia, previous hepatitis B virus infection, hypertension, diabetes mellitus, benign prostatic hyperplasia, acute myeloid leukemia | Fever, abdominal pain | Colonic diverticulitis | Cefepime and MTZ (10 days), SAM and MTZ (3 days) | Alive, discharged (228 days) |
| 7 | 30s/M | Epilepsy, under treatment for CRBSI with *Staphylococcus aureus* | Fever | Bacteremia of unknown origin | Cefazolin (5 days), SAM (12 days) | Alive, transferred (41 days) |
| 8 | 70s/M | Oral candida, adenocarcinoma (primary site unknown) | Septic shock, ascites, abdominal distension | Bowel obstruction | TZP (1 day)<br>TZP and fluconazole (1 day) | Death (1 day) |
| 9* | 60s/F | Rectal cancer | Abdominal pain, vomiting | Colon perforation, peritonitis | TZP (1 day), SAM (14 days), Hartmann's procedure | Alive, discharged (28 days) |
| 10* | 90s/F | Hypertension, dyslipidemia, osteoporosis, postoperative Mallory–Weiss syndrome, suspected aspiration pneumonia | Fever, tachycardia | Bowel obstruction | Cefotiam (4 days), SAM (15 days), intestinal resection | Alive, discharged (44 days) |
| 11* | 80s/F | Osteoporosis, dyslipidemia | Abdominal pain | Perforated appendicitis, peritonitis, intra-abdominal abscess | SAM (11 days), SAM and MTZ (8 days), appendectomy | Alive, discharged (19 days) |
| 12 | 70s/M | Cerebral infarction history, cholecystectomy history | Chills, fever, vomiting | Bacteremia of unknown origin | CPFX and MTZ (3 days), CRO and MTZ (10 days) | Alive (outpatient) |
| 13* | 80s/M | Hypertension, cholecystectomy history, rectal cancer | Abdominal pain | Colorectal perforation | SAM (10 days), TZP (12 days), Hartmann's procedure | Alive, discharged (53 days) |

AMP, ampicillin; CMZ, cefmetazole; CPFX, ciprofloxacin; transferred, transferred to another hospital.
\*In cases marked with an asterisk, multiple pathogens were detected as follows: 2, *A. onderdonkii*, *Bacteroides uniformis*, *E. coli*, and
*Pseudomonas nitroreducens*; 5, *A. putredinis* and *E. coli*; 9, *A. putredinis* and *Flavonifractor plautii*; 10, *A. indistinctus* and
*K. pneumoniae*; 11, *A. onderdonkii* and *Bacteroides salyersiae*; and 13, *A. finegoldii* and *A. ihumii*.
†Most patients received multiple antibiotics; the two used for the longest duration are shown.

### Clinical Outcomes

Of the 13 patients, 11 achieved clinical resolution following antimicrobial therapy, whereas the remaining 2 (cases 1 and 8) had a fatal outcome. The patient in fatal case 1 was a woman in her 80s with rectal cancer and liver metastases, who was admitted to the hospital for abdominal pain. Upon arrival at the hospital, the patient experienced septic shock. The patient received intravenous fluids, blood cultures were obtained, and TAZ and VCM were administered. Her condition briefly stabilized following the infusion, but subsequently deteriorated with the development of acidosis, hyperkalemia, and elevated lactic acid levels. The patient was diagnosed with colonic perforation and obstruction. The decision was made to transfer the patient to a hospital near her home for palliative care and to discontinue aggressive treatment. However, her condition deteriorated, and she died within the day of the transfer. After the patient died, *A. finegoldii* was detected in the blood culture obtained during an episode of septic shock.

The patient in fatal case 8 was a man in his 70s who was hospitalized for a comprehensive evaluation of adenocarcinoma of unknown primary origin. The patient was administered fluconazole for oral candidiasis. During hospitalization, the patient developed septic shock and was administered intravenous fluids. Blood cultures were obtained, and TAZ was administered. Although the patient initially stabilized, he later developed fecal emesis, likely due to intestinal obstruction. One day after the onset, the patient eventually died of septic shock. Blood cultures obtained during a septic shock episode revealed the presence of *A. onderdonkii*.

### Microbiological Characteristics

The microbiological characteristics of the *Alistipes* cases and strains are presented in Table 3. *Alistipes* strains formed small colonies on the Brucella HK agar medium (Fig. 1). MALDI-TOF MS identified 13 strains with species-level accuracy, including *A. finegoldii* (n = 4), *A. onderdonkii* (n = 4), *A. putredinis* (n = 3), and *A. indistinctus* (n = 2). Of the 13 strains, 16S rRNA and whole genome analyses were performed on 10 preserved strains (KML 24001–24010). 16S rRNA gene analysis showed that 10 of the preserved strains were 99.8–100% similar to the species identified by MALDI-TOF MS (Table S2).

**Fig 1.**
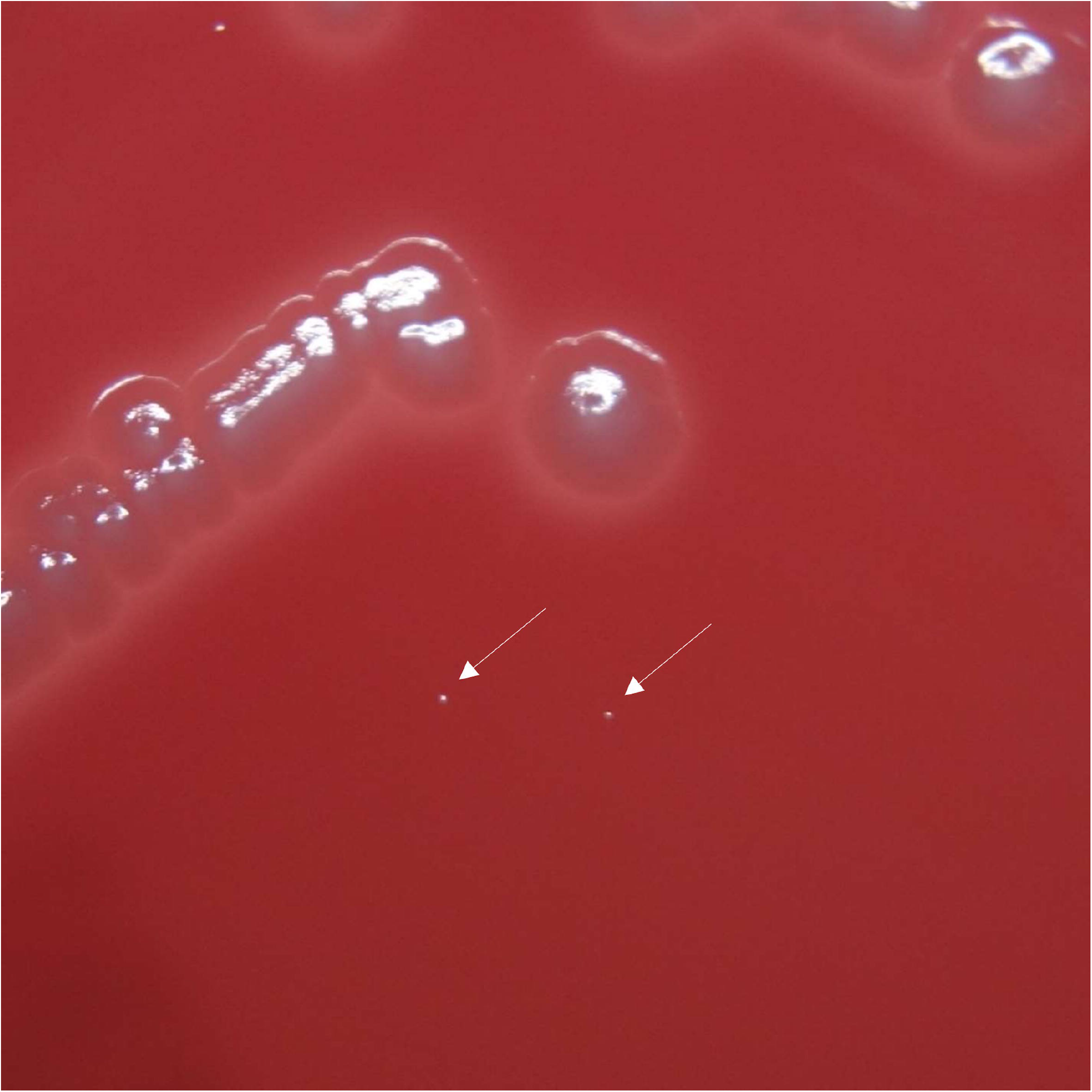
Small colonies of *Alistipes finegoldii* grown on agar medium in anaerobic culture for 48 h^*^. ^*^Figure 1 shows *A. finegoldii* (arrow mark) and *Escherichia coli* ATCC 25922 colonies grown on Brucella HK agar medium after 2 days of anaerobic culture.

**Table 3.** Identification results of MALDI-TOF MS, 16S rRNA analysis and whole genome analysis for the strains isolated from patients with *Alistipes* bacteremia

| Case no. | Strain name | MALDI-TOF MS |  | 16S rRNA analysis |  | Whole genome analysis |  |
| --- | --- | --- | --- | --- | --- | --- | --- |
|  |  | Proposed species | Score | Identified species | Similarity (%) | Identified species | ANI (%) |
| 1 | NA | *A. finegoldii* | 2.0 | NA | NA | NA | NA |
| 2 | NA | *A. onderdonkii* | 2.1 | NA | NA | NA | NA |
| 3 | NA | *A. finegoldii* | 2.1 | NA | NA | NA | NA |
| 4 | KML 24001 | *A. onderdonkii* | 2.3 | *A. onderdonkii* | 99.9 | *A. onderdonkii* | 98.3 |
| 5 | KML 24002 | *A. putredinis* | 2.1 | *A. putredinis* | 100 | *A. putredinis* | 99.9 |
| 6 | KML 24003 | *A. finegoldii* | 2.0 | *A. finegoldii* | 99.9 | *A. finegoldii* | 99.1 |
| 7 | KML 24004 | *A. indistinctus* | 2.0 | *A. indistinctus* | 100 | *A. indistinctus* | 99.1 |
| 8 | KML 24005 | *A. onderdonkii* | 2.1 | *A. onderdonkii* | 100 | *A. onderdonkii* | 99.6 |
| 9 | KML 24006 | *A. putredinis* | 2.1 | *A. putredinis* | 100 | *A. putredinis* | 99.7 |
| 10 | KML 24007 | *A. indistinctus* | 2.2 | *A. indistinctus* | 100 | *A. indistinctus* | 99.2 |
| 11 | KML 24008 | *A. onderdonkii* | 2.0 | *A. onderdonkii* | 100 | *A. onderdonkii* | 98.8 |
| 12 | KML 24009 | *A. putredinis* | 2.1 | *A. putredinis* | 99.9 | *A. putredinis* | 98.9 |
| 13 | KML 24010 | *A. finegoldii* | 2.3 | *A. finegoldii* | 99.8 | NA | NA |
| 13 | KML 24011 | No identification | NA | *A. ihumii* | 99.7 | NA | NA |

NA, not available.

The draft genomes of nine strains (KML 24001-24009) had contamination rates of < 1% and were used for further analysis (Table S3). One remaining strain (KML 24010) had a contamination rate of > 5% and was excluded from further analysis. The genome sizes of *A. finegoldii*, *A. onderdonkii*, *A. putredinis*, and *A. indistinctus* were 3.6 Mb, 3.3–3.6 Mb, 2.5–2.8 Mb, and 2.8–2.9 Mb, respectively (Table S4). The ANI results of KML 24001-24009 were in agreement with the results of MALDI-TOF MS, with an ANI of 99.8–100% (Table S5). DDH analysis for *A. onderdonkii* revealed that KML 24001 and KML 24005 showed DDH > 80% only for *A. onderdonkii* subsp. *vulgaris*, and KML 24008 showed DDH > 80% only for *A. onderdonkii* subsp. *onderdonkii*. The subspecies of each strain were identified (Table S6).

Strains with > 5% contamination were pure-cultured, and the two types of colonies grown were re-identified by MALDI-TOF MS. One strain (KML 24010) was identified as *A. finegoldii* by MALDI-TOF MS, and this strain showed 99.8% similarity to *A. finegoldii* by 16S rRNA gene analysis. Another strain (KML 24011) was not identified by MALDI-TOF MS and showed 99.7% similarity to *A. ihumii* AP11 (JX101692). The 16S rRNA gene sequence and draft genome data were deposited in GenBank via the DNA DataBank of Japan, and the accession numbers are given in Tables S2 and S4.

Reanalysis of the three *A. finegoldii* strains (3302398, 4401054, and Granada) isolated in previous reports yielded the following results: strain 3302398 showed 100% similarity to *A. dispar* 5CPEGH6 (accession no. AP019736), and strain 4401054 showed 99.0–100% similarity to *A. onderdonkii* subsp. *onderdonkii* DSM 19147 (ARFY01000017). Strain Granada showed 97.6% similarity with *A. finegoldii* DSM 17242 (CP003274); however, the species could not be confirmed.

### Antimicrobial Susceptibility and Resistance Genes

Antimicrobial susceptibility data were available for *Alistipes* 10 strains (Table 4). Low MIC_90_ values were observed for SAM (MIC_90_, 2 μg/mL), AMC (2 μg/mL), TZP (≤ 4 μg/mL), IPM (1 μg/mL), CLI (1 μg/mL), and MTZ (0.5 μg/mL). For CLI, only one *A. putredinis* strain showed an MIC of 2 μg/mL, and this strain possessed the *erm*(G) gene. The antibiotics with high MIC_90_ values were PEN (MIC_90_, > 1 μg/mL), CRO (> 32 μg/mL), FOX (32 μg/mL), MINO (8 μg/mL), and MXF (> 4 μg/mL). MICs were particularly high for PEN and MINO, with MIC_50_ values of > 1 μg/mL and 4 μg/mL, respectively. Seven strains with MICs ≥ 4 μg/mL for MINO harbored the *adeF* and *tet*(Q) genes. Two strains exhibited MICs of 32 μg/mL for FOX, and these strains were all *A. indistinctus*. Three strains showed MICs ≥ 4 μg/mL for MXF, all of which were *A. onderdonkii*.

**Table 4.**
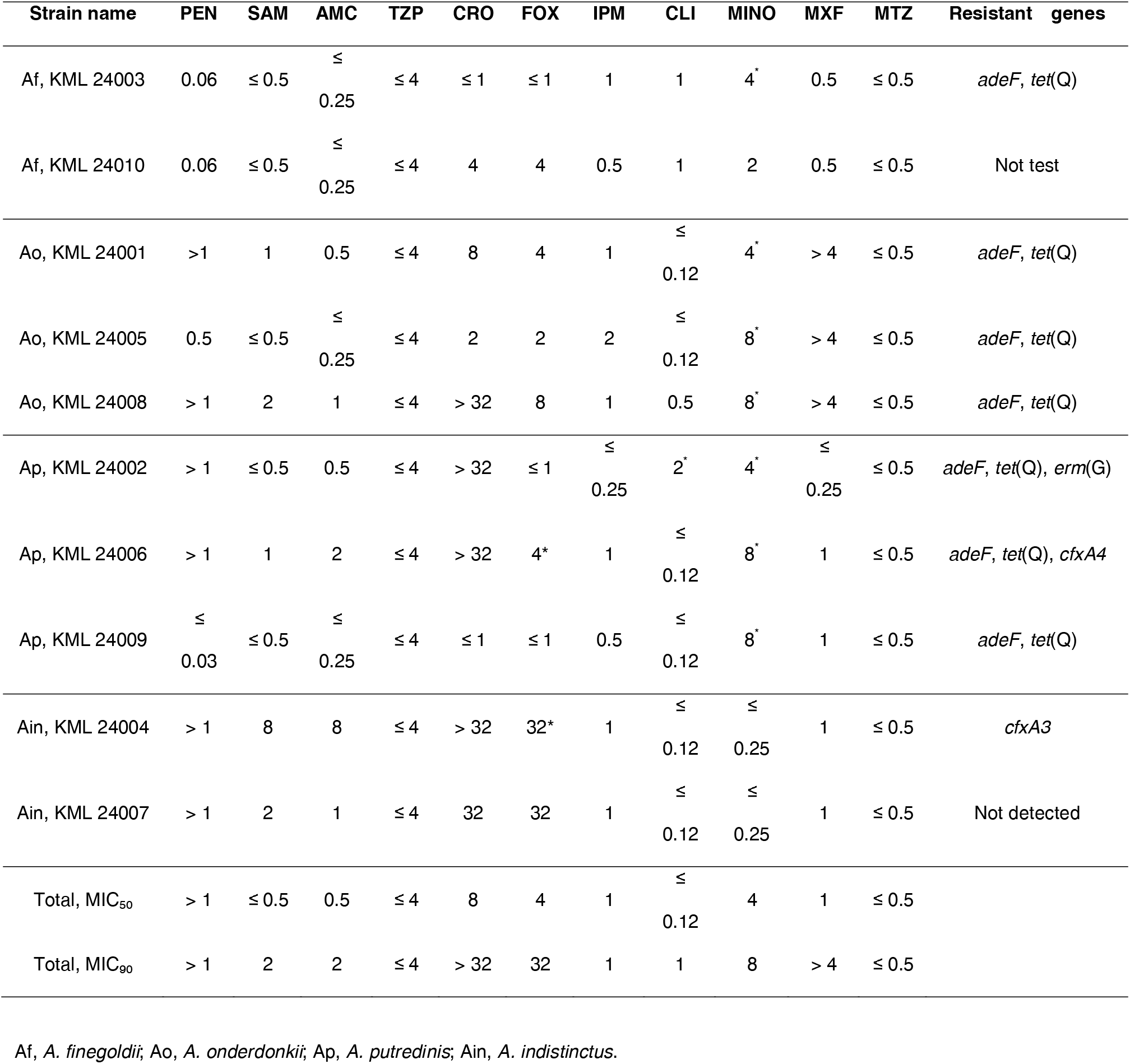
Minimum inhibitory concentration values (μg/mL) against antibiotics and antimicrobial-resistant genes of *Alistipes* strains

| Strain name | PEN | SAM | AMC | TZP | CRO | FOX | IPM | CLI | MINO | MXF | MTZ | Resistant genes |
| --- | --- | --- | --- | --- | --- | --- | --- | --- | --- | --- | --- | --- |
| Af, KML 24003 | 0.06 | ≤ 0.5 | ≤<br>0.25 | ≤ 4 | ≤ 1 | ≤ 1 | 1 | 1 | 4<sup>+</sup> | 0.5 | ≤ 0.5 | *adeF*, *tet(Q)* |
| Af, KML 24010 | 0.06 | ≤ 0.5 | ≤<br>0.25 | ≤ 4 | 4 | 4 | 0.5 | 1 | 2 | 0.5 | ≤ 0.5 | Not test |
| Ao, KML 24001 | > 1 | 1 | 0.5 | ≤ 4 | 8 | 4 | 1 | ≤<br>0.12 | 4<sup>+</sup> | > 4 | ≤ 0.5 | *adeF*, *tet(Q)* |
| Ao, KML 24005 | 0.5 | ≤ 0.5 | ≤<br>0.25 | ≤ 4 | 2 | 2 | 2 | ≤<br>0.12 | 8<sup>+</sup> | > 4 | ≤ 0.5 | *adeF*, *tet(Q)* |
| Ao, KML 24008 | > 1 | 2 | 1 | ≤ 4 | > 32 | 8 | 1 | 0.5 | 8<sup>+</sup> | > 4 | ≤ 0.5 | *adeF*, *tet(Q)* |
| Ap, KML 24002 | > 1 | ≤ 0.5 | 0.5 | ≤ 4 | > 32 | ≤ 1 | ≤<br>0.25 | 2<sup>+</sup> | 4<sup>+</sup> | ≤<br>0.25 | ≤ 0.5 | *adeF*, *tet(Q)*, *erm(G)* |
| Ap, KML 24006 | > 1 | 1 | 2 | ≤ 4 | > 32 | 4<sup>+</sup> | 1 | ≤<br>0.12 | 8<sup>+</sup> | 1 | ≤ 0.5 | *adeF*, *tet(Q)*, *cfxA4* |
| Ap, KML 24009 | ≤<br>0.03 | ≤ 0.5 | ≤<br>0.25 | ≤ 4 | ≤ 1 | ≤ 1 | 0.5 | ≤<br>0.12 | 8<sup>+</sup> | 1 | ≤ 0.5 | *adeF*, *tet(Q)* |
| Ain, KML 24004 | > 1 | 8 | 8 | ≤ 4 | > 32 | 32<sup>*</sup> | 1 | ≤<br>0.12 | ≤<br>0.25 | 1 | ≤ 0.5 | *cfxA3* |
| Ain, KML 24007 | > 1 | 2 | 1 | ≤ 4 | 32 | 32 | 1 | ≤<br>0.12 | ≤<br>0.25 | 1 | ≤ 0.5 | Not detected |
| Total, MIC<sub>50</sub> | > 1 | ≤ 0.5 | 0.5 | ≤ 4 | 8 | 4 | 1 | ≤<br>0.12 | 4 | 1 | ≤ 0.5 |  |
| Total, MIC<sub>90</sub> | > 1 | 2 | 2 | ≤ 4 | > 32 | 32 | 1 | 1 | 8 | > 4 | ≤ 0.5 |  |

Af, *A. finegoldii*; Ao, *A. onderdonkii*; Ap, *A. putredinis*; Ain, *A. indistinctus*.

## Discussion

In this study, we identified 13 cases of *Alistipes* bacteremia at a single institution in Japan. Three main characteristics were observed in patients with *Alistipes* bacteremia. First, most patients were elderly or had comorbid conditions, with 7 of the 13 patients having either solid tumors or hematological malignancies. Second, general cases were associated with gastrointestinal diseases, and all patients were treated for true bacteremia. Third, some *Alistipes* strains had high MICs against β-lactams, tetracyclines, and quinolones, indicating the presence of antibiotic-resistant genes.

Anaerobic bacteria can cause infections in almost any part of the body, including severe bloodstream infections (*25*). In our study, 13 cases of bacteremia were caused by *A. finegoldii*, *A. putredinis*, *A. onderdonkii* subsp. *onderdonkii*, *A. onderdonkii* subsp. *vulgaris*, *A. indistinctus*, and *A. ihumii*. There have only been three *A. finegoldii* cases of *Alistipes* bacteremia reported to date (*12*, *13*), and Tyrrel et al. reported one of these strains (strain 4401054) to be *A. onderdonkii* (*7*). Our data support the report by Tyrrel et al., who showed that another strain (strain 3302398) was *A. dispar*. Slow growth and difficulty in identification probably contribute to the underestimation of *Alistipes* bacteremia. Compared with other bacteria, such as *E. coli* and *Bacteroides* species, *Alistipes* grows more slowly and forms significantly small colonies. Additionally, *Alistipes* is difficult to identify to by biochemical characteristics. However, the advent of MALDI-TOF MS has improved the ability to accurately identify anaerobes, including lesser known pathogens (*25*, *26*). Our study showed that MALDI-TOF MS could dentify *Alistipes* strains with high accuracy, in agreement with 16S rRNA gene analysis and whole genome sequencing. Although *Alistipes* bacteremia has not been recognized until recently, the number of cases is expected to increase in the future.

*Alistipes* species have been isolated from feces (*3*, *6*, *27*), and *A. putredinis* is commonly found in the gut microbiota (*28*). In previous cases of *Alistipes* bacteremia, two patients had colon cancer (*12*), and the other had intestinal perforation and peritonitis (*13*). Similarly, in our study, most patients (11/13) had underlying gastrointestinal symptoms. These findings suggest that *Alistipes* translocates from the gastrointestinal tract into the bloodstream. In our cohort, all patients were treated with antibiotics for true bacteremia, and 11 recovered. Two patients who died were aged ≥ 70 years, had solid tumors, and developed sepsis. These patients had monomicrobial bacteremia caused by *Alistipes* and were treated with antibiotics to which the isolates were susceptible. The fatal outcomes were likely due to the complications of sepsis and the effects of underlying malignancies.

*Alistipes* strains showed different antimicrobial susceptibilities depending on the species and strain. All the strains showed low MIC values for BLBLIs, IPM, and MNZ. Consistent with our findings, previously reported strains were also susceptible to SAM (*7*), TZP (*13*), and MNZ (*3*, *7*, *13*). In our cohort, 12 patients were treated with BLBLIs and 10 recovered. In previously reported cases of *Alistipes* bacteremia, three patients were successfully treated with combinations of AMC and ciprofloxacin, AMC and amikacin, or TZP and MTZ (*12*, *13*). BLBLIs and MNZ are commonly used for anaerobic infections (*29*) and can be an option for the treatment of *Alistipes* bacteremia.

Conversely, several *Alistipes* strains were resistant to PEN, CRO, MINO, MXF, and FOX. Low susceptibility to MXF and FOX was observed only in *A. onderdonkii* and *A. indistinctus*. Moreover, our data revealed the presence of antibiotic-resistant genes, such as *adeF*, *tet*(Q), *cfxA3*, *cfxA4*, and *erm*(G). Consistent with our findings, previous reports have indicated resistance to PEN (*11–13*), tetracyclines (*3*), and MXF (*11*) in *Alistipes* strains. Although breakpoints for *Alistipes* have not been established by the Clinical and Laboratory Standards Institute (*30*), it may be prudent to avoid using antibiotics that have been observed to have low susceptibility for the treatment of *Alistipes* bacteremia.

The limitations of this study are its single-center design and the limited number of cases. As this was a single-center study, the generalizability of patient and strain characteristics is limited. The limited number of cases for each strain prevented statistical comparisons of patient backgrounds and antimicrobial susceptibilities between the species. Therefore, future studies should include a wider range of regions and institutions. Moreover, some strains were not preserved, which limited the validation of the identification tests and antimicrobial susceptibility measurements for these non-preserved strains. Furthermore, discrepancies between phenotypic and genotypic antimicrobial susceptibilities were observed, and not all resistance mechanisms could be elucidated. Our study highlights the need for further evaluation, including mutational analysis and efflux pump assessment, to fully understand the resistance mechanisms.

In conclusion, 13 cases of *Alistipes* bacteremia were observed over a 7-year period at a tertiary care center in Japan. Most patients were elderly and had comorbid conditions involving solid tumors or hematological malignancies. The most common symptoms were gastrointestinal, and two of the patients died of sepsis. MALDI-TOF MS will facilitate the identification of *Alistipes* strains, and reports of *Alistipes* bacteremia are predicted to increase.

## About the Author

Naoki Watanabe is a clinical laboratory scientist specializing in infectious disease at Kameda Medical Center in Kamogawa, Japan. His primary interests include infectious diseases, microbiological testing, and antimicrobial stewardship.

## Data Availability

All data produced in the present study are available upon reasonable request to the authors.

## Acknowledgment

We thank the clinical laboratory scientist at Kameda Medical Center for their assistance with microbiological testing.

## REFERENCES

1. Rautio M, Lönnroth M, Saxén H, Nikku R, Väisänen ML, Finegold SM, et al. Characteristics of an unusual anaerobic pigmented gram-negative rod isolated from normal and inflamed appendices. Clin Infect Dis. 1997 Sep;25 Suppl 2:S107–10. 10.1086/516210

2. Rautio M, Saxén H, Siitonen A, Nikku R, Jousimies-Somer H. Bacteriology of histopathologically defined appendicitis in children. Pediatr Infect Dis J. 2000 Nov;19(11):1078–83. 10.1097/00006454-200011000-00010

3. Rautio M, Eerola E, Väisänen-Tunkelrott ML, Molitoris D, Lawson P, Collins MD, et al. Reclassification of *Bacteroides* putredinis (Weinberg et al., 1937) in a new genus *Alistipes* gen. nov., as *Alistipes* putredinis comb. nov., and description of *A. finegoldii* sp. nov., from human sources. Syst Appl Microbiol. 2003 Jun;26(2):182–8. 10.1078/072320203322346029

4. Song Y, Könönen E, Rautio M, Liu C, Bryk A, Eerola E, et al. *Alistipes onderdonkii* sp. nov. and *Alistipes* shahii sp. nov., of human origin. Int J Syst Evol Microbiol. 2006 Aug;56(Pt 8):1985–90. 10.1099/ijs.0.64318-0

5. Simmon KE, Mirrett S, Reller LB, Petti CA. Genotypic diversity of anaerobic isolates from bloodstream infections. J Clin Microbiol. 2008 May;46(5):1596–601. 10.1128/JCM.02469-07

6. Nagai F, Morotomi M, Watanabe Y, Sakon H, Tanaka R. *A. indistinctus* sp. nov. and Odoribacter laneus sp. nov., common members of the human intestinal microbiota isolated from faeces. Int J Syst Evol Microbiol. 2010 Jun;60(Pt 6):1296–302. 10.1099/ijs.0.014571-0

7. Tyrrell KL, Warren YA, Citron DM, Goldstein EJC. Re-assessment of phenotypic identifications of *Bacteroides* putredinis to *Alistipes* species using molecular methods. Anaerobe. 2011 Jun;17(3):130–4. 10.1016/j.anaerobe.2011.04.002

8. Dziarski R, Park SY, Kashyap DR, Dowd SE, Gupta D. Pglyrp-Regulated Gut Microflora Prevotella falsenii, ParaBacteroides distasonis and Bacteroides eggerthii Enhance and A. finegoldii Attenuates Colitis in Mice. PLoS One. 2016 Jan 4;11(1):e0146162. 10.1371/journal.pone.0146162

9. Naseribafrouei A, Hestad K, Avershina E, Sekelja M, Linløkken A, Wilson R, et al. Correlation between the human fecal microbiota and depression. Neurogastroenterol Motil. 2014 Aug;26(8):1155–62. 10.1111/nmo.12378

10. Lassmann B, Gustafson DR, Wood CM, Rosenblatt JE. Reemergence of anaerobic bacteremia. Clin Infect Dis. 2007 Apr 1;44(7):895–900. 10.1086/512197

11. Cobo F, Foronda C, Pérez-Carrasco V, Martin-Hita L, García-Salcedo JA, Navarro-Marí JM. First description of abdominal infection due to *Alistipes* onderdonkii. Anaerobe. 2020 Dec;66:102283. 10.1016/j.anaerobe.2020.102283

12. Fenner L, Roux V, Ananian P, Raoult D. *A. finegoldii* in blood cultures from colon cancer patients. Emerg Infect Dis. 2007 Aug;13(8):1260–2. 10.3201/eid1308.060662

13. Cobo F, Franco-Acosta A, Lara-Oya A, Pérez-Carrasco V. Bacteremia caused by *A. finegoldii* in a patient with peritonitis. Enferm Infecc Microbiol Clin. 2023 Feb;41(2):131–2. 10.1016/j.eimce.2022.11.002

14. Salonen JH, Eerola E, Meurman O. Clinical significance and outcome of anaerobic bacteremia. Clin Infect Dis. 1998 Jun;26(6):1413–7. 10.1086/516355

15. 15. Lane DJ. 16S/23S rRNA sequencing. Nucleic acid techniques in bacterial systematics. 1991; https://cir.nii.ac.jp/crid/1570009750603612672

16. Jaric M, Segal J, Silva-Herzog E, Schneper L, Mathee K, Narasimhan G. Better primer design for metagenomics applications by increasing taxonomic distinguishability. BMC Proc. 2013 Dec 20;7(Suppl 7):S4. 10.1186/1753-6561-7-S7-S4

17. Watanabe N, Watari T, Otsuka Y, Hosokawa N, Yamagata K, Fujioka M. Clinical and microbiological characteristics of Ruminococcus gnavus bacteremia and intra-abdominal infection. Anaerobe. 2024 Jan 9;102818. 10.1016/j.anaerobe.2024.102818

18. Yoon SH, Ha SM, Kwon S, Lim J, Kim Y, Seo H, et al. Introducing EzBioCloud: a taxonomically united database of 16S rRNA gene sequences and whole-genome assemblies. Int J Syst Evol Microbiol. 2017 May;67(5):1613–7. 10.1099/ijsem.0.001755

19. Parks DH, Imelfort M, Skennerton CT, Hugenholtz P, Tyson GW. CheckM: assessing the quality of microbial genomes recovered from isolates, single cells, and metagenomes. Genome Res. 2015 Jul;25(7):1043–55. 10.1101/gr.186072.114

20. Tanizawa Y, Fujisawa T, Nakamura Y. DFAST: a flexible prokaryotic genome annotation pipeline for faster genome publication. Bioinformatics. 2018 Mar 15;34(6):1037–9. 10.1093/bioinformatics/btx713

21. Meier-Kolthoff JP, Carbasse JS, Peinado-Olarte RL, Göker M. TYGS and LPSN: a database tandem for fast and reliable genome-based classification and nomenclature of prokaryotes. Nucleic Acids Res. 2022 Jan 7;50(D1):D801–7. 10.1093/nar/gkab902

22. Meier-Kolthoff JP, Hahnke RL, Petersen J, Scheuner C, Michael V, Fiebig A, et al. Complete genome sequence of DSM 30083(T), the type strain (U5/41(T)) of *Escherichia coli*, and a proposal for delineating subspecies in microbial taxonomy. Stand Genomic Sci. 2014 Dec 8;9:2. 10.1186/1944-3277-9-2

23. Alcock BP, Huynh W, Chalil R, Smith KW, Raphenya AR, Wlodarski MA, et al. CARD 2023: expanded curation, support for machine learning, and resistome prediction at the Comprehensive Antibiotic Resistance Database. Nucleic Acids Res. 2023 Jan 6;51(D1):D690–9. 10.1093/nar/gkac920

24. Kanda Y. Investigation of the freely available easy-to-use software “EZR” for medical statistics. Bone Marrow Transplant. 2013 Mar;48(3):452–8. 10.1038/bmt.2012.244

25. Nagy E, Boyanova L, Justesen US, ESCMID Study Group of Anaerobic Infections. How to isolate, identify and determine antimicrobial susceptibility of anaerobic bacteria in routine laboratories. Clin Microbiol Infect. 2018 Nov;24(11):1139–48. 10.1016/j.cmi.2018.02.008

26. Justesen Ulrik Stenz, Holm Anette, Knudsen Elisa, Andersen Line Bisgaard, Jensen Thøger Gorm, Kemp Michael, et al. Species Identification of Clinical Isolates of Anaerobic Bacteria: a Comparison of Two Matrix-Assisted Laser Desorption Ionization–Time of Flight Mass Spectrometry Systems. J Clin Microbiol. 2020 Dec 21;49(12):4314–8. 10.1128/jcm.05788-11

27. Sakamoto M, Ikeyama N, Ogata Y, Suda W, Iino T, Hattori M, et al. *Alistipes* communis sp. nov., A. dispar sp. nov. and Alistipes onderdonkii subsp. vulgaris subsp. nov., isolated from human faeces, and creation of Alistipes onderdonkii subsp. onderdonkii subsp. nov. Int J Syst Evol Microbiol. 2020 Jan;70(1):473–80. 10.1099/ijsem.0.003778

28. Qin J, Li R, Raes J, Arumugam M, Burgdorf KS, Manichanh C, et al. A human gut microbial gene catalogue established by metagenomic sequencing. Nature. 2010 Mar4;464(7285):59–65. 10.1038/nature08821

29. Brook I, Wexler HM, Goldstein EJC. Antianaerobic antimicrobials: spectrum and susceptibility testing. Clin Microbiol Rev. 2013 Jul;26(3):526–46. 10.1128/CMR.00086-12

30. Clinical and Laboratory Standards Institute - Performance Standards for Antimicrobial Susceptibility Testing, 34th Edition. [cited 2024 July 1]. https://em100.edaptivedocs.net

